# Epigenetic Element-Based Transcriptome-Wide Association Study Identifies Novel Genes for Bipolar Disorder

**DOI:** 10.1101/2020.07.23.20161174

**Authors:** Shi Yao, Jing-Miao Ding, Hao Wu, Ruo-Han Hao, Yu Rong, Xin Ke, Jing Guo, Shan-Shan Dong, Tie-Lin Yang, Yan Guo

## Abstract

Since the bipolar disorder (BD) signals identified by genome-wide association study (GWAS) often reside in the non-coding regions, understanding the biological relevance of these genetic loci has proven to be complicated. Transcriptome-wide association studies (TWAS) providing a powerful approach to identify novel disease risk genes and uncover possible causal genes at loci identified previously by GWAS. However, these methods did not consider the importance of epigenetic regulation in gene expression. Here, we developed a novel epigenetic element-based transcriptome-wide association study (ETWAS) that tests the effects of genetic variants on gene expression levels with the epigenetic features as prior and further mediates the association between predicted expression and BD. We conducted an ETWAS consisting of 20,352 cases and 31,358 controls and identified 44 transcriptome-wide significant hits. We found 14 conditionally independent genes, and 11 did not previously implicate with BD, which is regarded as novel candidate genes, such as *ASB16* in the cerebellar hemisphere (*P* = 9.29×10^−8^). We demonstrated that several genome-wide significant signals from the BD GWAS driven by genetically regulated expression, and conditioning of *NEK4* explaining 90.1% of the GWAS signal. Additionally, ETWAS identified genes could explain heritability beyond that explained by GWAS-associated SNPs (*P* = 0.019). By querying the SNPs in the final model of identified genes in phenome databases, we identified several phenotypes previously associated with BD, such as schizophrenia and depression. In conclusion, ETWAS is a powerful method, and we identified several novel candidate genes associated with BD.

## Introduction

Bipolar disorder (BD) is a severe neuropsychiatric disorder characterized by recurrent episodes of depression and mania that affect thought, perception, emotion, and social behavior. BD has a lifetime prevalence of 1-2%, and the World Health Organization ranks BD among the most significant contributors to the global burden of disease ^1^. Based on twin studies, the narrow-sense heritability of BD has estimated to be over 70% ^2, 3^. Genome-wide association study (GWAS) has seen great strides and invaluable utilities in revealing initial insights into BD’s genetic architecture. Recently, a large-scale GWAS identified 30 loci that were significantly associated with BD ^4^. Despite the significant success of GWAS in delineating elements that contribute to the genetic architecture of psychiatric disorders, only a small fraction of this heritability is explained by associated loci ^4^, leaving a substantial proportion of genetic risk factors uncharacterized.

Most of the identified variants mapped through GWAS reside in non-coding regions of the genome ^5^, that may be involved in modulating gene regulatory programs ^5-9^. Recent mechanistic studies have demonstrated that GWAS-identified variants located in the active chromatin regions more frequently and highly-enriched with expression quantitative trait loci (eQTL) ^10, 11^. Moreover, most common risk variants identified to date are only associated with diseases with modest effect sizes, and many risk variants have not identified via a typical GWAS, even with a large sample size ^12^. Transcriptome-wide association study (TWAS) that systematically investigates the association of genetically predicted gene expression with disease risk, providing a powerful approach to identify novel disease risk genes and uncover possible causal genes at loci identified previously by GWAS ^13-17^.

Nevertheless, gene expression highly regulated in many steps, including transcriptional regulation, splicing, end modification, export, and degradation. Transcriptional regulation of DNA into mRNA can occur on both genetic and epigenetic levels. The epigenetic regulation alters the accessibility of DNA to transcription factors by chemical modification of chromatin. For example, several post-translational modifications occur on the histones that could change chromatin structure and function ^18^, making it accessible or vice versa to transcription factors. Functional class quantification in 11 diseases from the Wellcome Trust Case Control Consortium, including BD, showed that 80% of the common variants that contribute to phenotype variability could attribute to DNase I hypersensitivity sites ^19^. They are likely to regulate chromatin accessibility and transcription, further highlighting the importance of transcript regulation in the epigenetic level. Histone modifications are involved in both activation and repression of transcription ^20^ and further linked to diseases ^21^. Researchers have developed a growing body of computational methods to predict gene expression from histone modification signals of chromatin structure ^22-25^. Thus, integrating epigenetic features is essential for the prediction of gene expression besides genetic variants.

In this study, we set out to develop a four-step quantitative pipeline named epigenetic element-based transcriptome-wide association studies (ETWAS), based on the interpretation of epigenetic element, genotype, gene expression, and phenotype. We used ETWAS to investigate the association between gene expression and BD risk using the largest BD cohort currently available (as of 2020); the cohort consists of 20,352 BD cases and 31,358 controls from Europe. We found that ETWAS outperformed than original methods, and we identified 14 conditionally independent genes associated with BD risk in 13 brain tissues. We additionally identified 11 genes that not previously implicated with BD.

## Methods

### Data Resources

#### RNA sequencing data sets

We used transcriptome and high-density genotyping data of European decedent from the Genotype-Tissue Expression (GTEx) study Pilot Project V8 (dbGap accession: phs000424.v8.p2) ^11^ to establish gene expression prediction models (Supplementary Methods). We also obtained freely available RNA-seq data from 358 European lymphoblastoid cell lines produced by the Genetic European Variation in Health and Disease ^26^ (Geuvadis) at https://www.ebi.ac.uk/ as the validation data set to test the prediction models generated in the GTEx whole blood. The tissue abbreviations, sample sizes are listed in Supplementary Table 1.

#### Epigenetic elements

The chromatin states of relevant tissues were downloaded from the Roadmap Epigenome Project ^27^. We briefly utilized the 15-state model and grouped them into four categories: *promoter, enhancer, transcription*, and *others*. Individually, *TssA* and *TssAFlnk* are considered to be a *promoter*; *TxFlnk, Tx*, and *TxWk* are considered to be a *transcription*; *EnhG* and *Enh* are considered to be an *enhancer*, and the rest are classified as the *others*. Freely available transcription factor binding sites (TFBS) were obtained from the Encyclopedia of DNA Elements (ENCODE) ^28^, and the DNase I hypersensitive sites (DHS) were obtained from the Roadmap Epigenome Project. The URLs of all epigenetic data are listed in Supplementary Table 2.

#### GWAS summary statistics

We used the most recent summary statistics of the Psychiatric Genomics Consortium (PGC) Bipolar Disorder Working Group, comprising 20,352 BD cases and 31,358 controls of European descent (Supplementary Table 3). Details on participant ascertainment and quality control were previously reported by Eli et al. ^4^ (Supplementary Methods). We evaluated ETWAS’s performance by identifying significantly associated genes in an earlier reported summary statistic that did not overlap a genome-wide significant SNP, and looking for newly genome-wide significant SNPs in the expanded study. The earlier reported summary data is the first large collaborative BD GWAS by the same group in PGC ^29^, comprised of 7,481 BD patients and 9,250 controls of European descent (Supplementary Table 3).

### ETWAS Framework

Our current study based on the premise that gene expression is heritable ^30^. Considering the high heritability genes typically enriched for trait associations ^15^, we only focused on the highly heritable genes in further analyses (Supplementary Methods).

Our prediction framework includes four main sections which act sequentially (Figure 1, Supplementary Methods). First, for each gene, we divide the SNPs within 1 Mb of the transcription start/end site of the gene into multiple SNP sets according to the eQTL *P*-value and epigenetic annotations. We construct multiple elastic net and lasso models with the SNPs in each SNP set using the initial reference data. Second, we evaluate the prediction performance of each SNP set using tenfold cross-validation R^2^ between the predicted and observed expression, and the SNP set with the highest mean R^2^ is selected as the best model. Third, we construct the final prediction model with the parameters of the best SNP set using all the samples in the reference data. We estimate the associations between predicted expressions and traits with the combination of SNP-trait effect sizes while accounting for linkage disequilibrium among SNPs.

**Figure 1.**
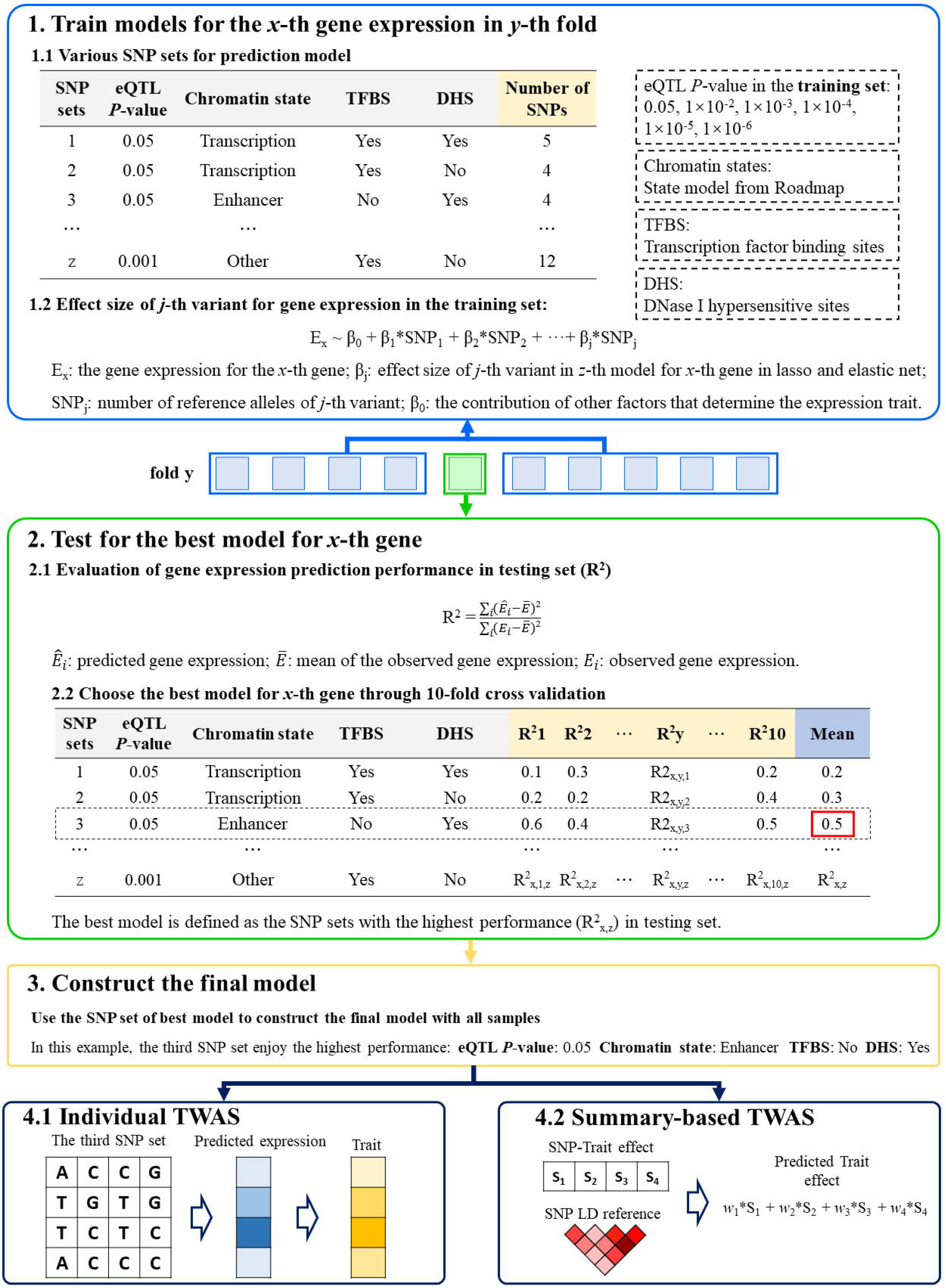
Schematic of ETWAS approach.

### Evaluating Prediction Accuracy

The within-study prediction was evaluated in the whole blood and 13 brain tissues of the highly heritable genes. The accuracy was evaluated using tenfold cross-validation *R*^*2*^ between predicted and true expression. We then evaluated the prediction models on an independent cohort. Cross-study prediction accuracy was measured by using weights derived from the best models with all whole blood samples to predict gene expression levels in the Geuvadis dataset.

We compared the cross-validation *R*^*2*^ between ETWAS and recent work in parallel to ours, which imputed expression using only genetic variants with different models, such as lasso, and elastic net ^13, 16^. Briefly, lasso uses an L1 penalty as a variable selection method to select a sparse of predictors ^31^. In contrast, the elastic net linearly combines the L1 and L2 penalties of lasso and ridge regression to perform variable selection ^32^. We used the R package *glmnet* to implement lasso and elastic net (α = 0.5).

### Transcriptomic Imputation

ETWAS was done using gene expression prediction models derived above in 13 brain tissues. A strict Bonferroni-corrected study-wise threshold was used: *P* = 2.55×10^−6^ (0.05/19,632, the total number of highly heritable genes across tissues). FUSION (http://gusevlab.org/projects/fusion/) was used to conduct the transcriptome-wide association testing. The 1000 Genomes v3 LD panel was used for the ETWAS.

### Conditionally Testing GWAS Signals

To determine how much GWAS signal remains after the expression association from ETWAS is removed, conditional and joint testing was done for genome-wide Bonferroni-corrected ETWAS genes via FUSION. Each BD GWAS SNP associations were conditioned on the joint gene model one SNP at a time. The overlapping genes were set to be in the range of 500 KB around each SNP, and the defined regions included only the transcribed region of the genes. The FUSION tool was used to perform the conditional and joint analyses with the *cis*-genetic component of expression we generated.

### Functional Relevance of Conditionally Independent Genes

We examined whether the gene or transcript expression of the identified genes were differentially expressed between BD patients and controls and further used the Mouse Genome Informatics (MGI) ^33^ to check whether the conditionally independent genes are relevant to BD (Supplementary Methods).

### Partitioned Heritability

To investigate whether the ETWAS identified genes could explain additional heritability, partitioned heritability of BD was estimated using LD score regression (LDSC) following previously described methodology ^34^ (Supplementary Methods).

### Enrichment Analyses

We performed the Genetic Association Database (GAD) disease enrichment analyses, GWAS catalog enrichment analyses, and drug targets enrichment analyses (Supplementary Methods) to demonstrate ETWAS’s ability to identify BD-related genes.

### Phenome-wide Association Studies

To identify phenotypes that may be associated or co-morbid with BD, we conducted a phenome-wide association study (pheWAS) for each SNP in the final model of the identified genes (Supplementary Methods).

## Results

### Model Generation and Evaluation

#### Heritability of Gene Expression

The mean heritability of gene expression is 0.016 for all protein-coding genes in whole blood, and 0.04 in brain tissues range from 0.031 to 0.049 in different tissues (Supplementary Table 1). We identified 2,239 high heritability genes in whole blood and 19,632 highly heritable gene-tissue pairs of 9,492 genes in 13 brain tissues. Among the heritable genes in the brain, we found that almost half of them (5,155/9,492) are significant only in one brain tissue (Supplementary Figure 1). We observed a high correlation of gene expression heritability in different brain tissues, with a correlation coefficient ranging from 0.40 to 0.79 (Supplementary Figure 2).

#### Epigenetic Annotation Improve the Performance of Gene Expression Prediction

The prediction performance of gene expression prediction is better in highly heritable genes (Supplementary Figure 3), which can be supplementary to prove the appropriateness of focusing on high heritability genes. We evaluated whether the expression of highly heritable genes could be accurately imputed in 13 brain tissues from genotype with the epigenetic elements as prior. We noted that the cross-validation performance significantly increased with the number of active annotations increasing in 11 tissues, and the genes with at least two active annotations 1.01× to 1.19× outperformed than the genes with 0 active annotations (Figure 2A, Supplementary Figure 4). We identified a negative correlation between cross-validation *R*^*2*^ and SNP number in the best models (Figure 2B, Supplementary Table 4), which consistent with the sparsity of the local architecture of gene expression and a handful of genetic variants seem to contribute to the variability in gene expression ^35^. We demonstrated that the best SNP sets for predicting gene expression significantly enriched in active epigenetic elements by comparing the epigenetic annotation frequency between the best SNP sets and all variants used (Pearson’s chi-squared test, *P* < 0.01) (Figure 2C, Supplementary Figure 5). Specifically, the best SNP sets are distributed in the active HMM annotation at a frequency of 1.32-1.45 times that of all variants, while distributed in the TFBS region and DHS region at a frequency of 2.12-2.43 and 1.70-2.40 times that of all variants. We further evaluated ETWAS’s performance and compared them with recent work in parallel to ours via cross-validation. On average, our pipeline attained slightly better performance than using lasso, elastic net, and top SNP (Figure 2D, Supplementary Figure 6). For ETWAS, the average tenfold cross-validated prediction *R*^*2*^ value ranging from 0.11 to 0.16 in different brain tissues (Supplementary Table 4).

**Figure 2.**
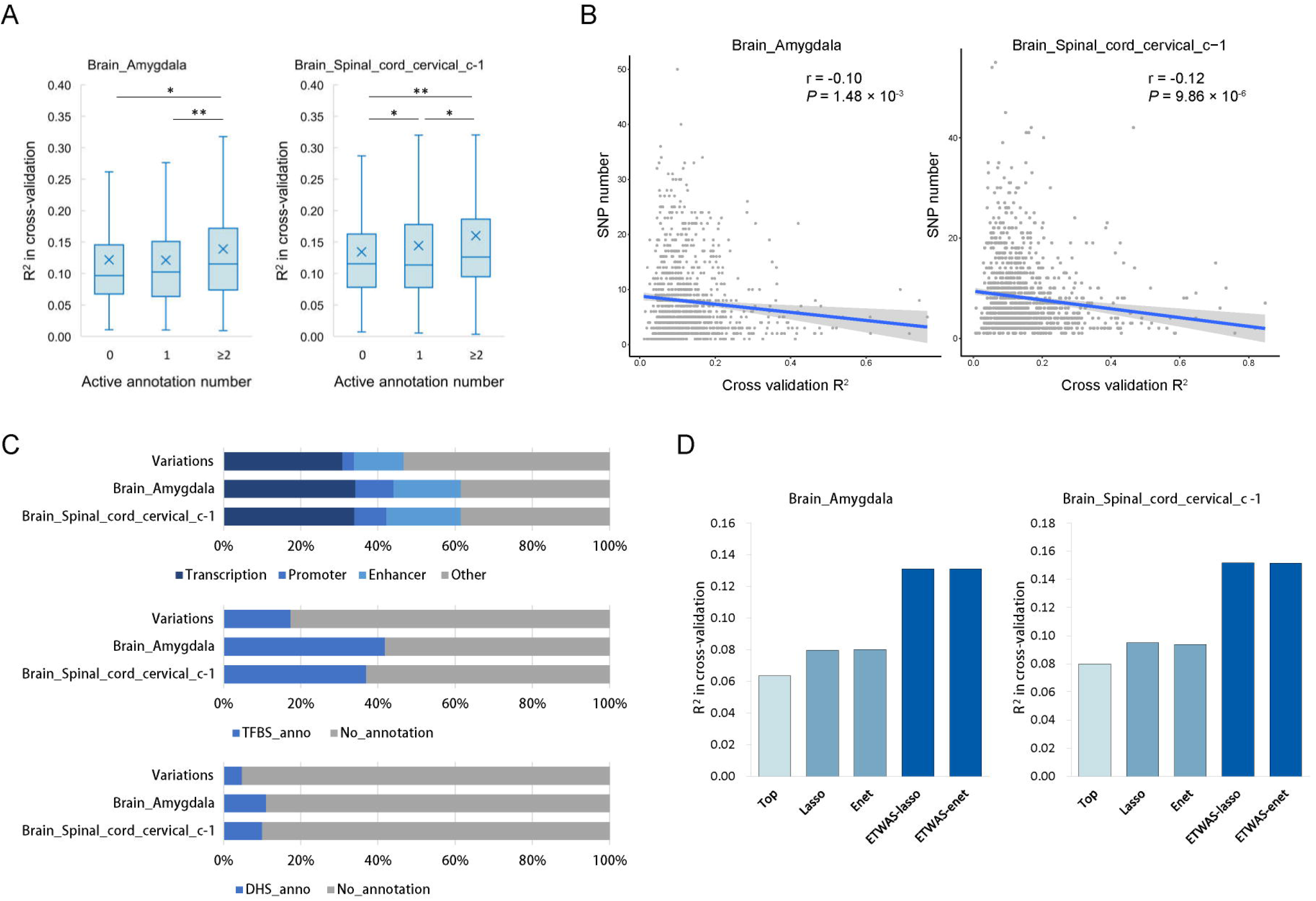
Epigenetic data improved the performance of gene expression prediction in brain tissues. (A) The cross-validation *R*^*2*^ of the best prediction models were sorted according to the active annotation number and grouped into three categories: 0, 1, ≥2. One asterisk (*) indicates *P*-value smaller than 0.05 (*P* < 0.05), two asterisks (**) indicates *P*-value smaller than 0.01 (*P* < 0.01). (B) The correlation between the prediction performance and SNP number used in the best model. The x-axis represents cross-validation R^2^ and the y-axis represents the SNP number of the best model. (C) The epigenetic annotation distributions of the SNPs used for all genes and the best ETWAS models in two brain tissues. (D) Accuracy of individual-level expression imputation models. Accuracy was estimated using cross-validation R^2^ between predicted and true expression. Bars show the mean estimate across 2 brain tissues and five methods: Top, Lasso, Enet, ETWAS.lasso and ETWAS.enet.

#### ETWAS Performance in a Separate Cohort

We also tested the prediction models on separate cohorts. We used prediction models trained in GTEx blood to compare predicted and observed expression in Geuvadis. The average prediction *R*^*2*^ is 0.034. The top six genes with the highest performance from this analysis are illustrated in Supplementary Figure 7, proving a comparison of the predicted and observed expression. Among these genes, the *CHURC1* trained in GTEx performed best, and the *R*^*2*^ between predicted and observed expression levels in Geuvadis is 0.80. A quantile-quantile plot showed expected and observed *R*^*2*^ from ETWAS is given (Supplementary Figure 8). We found a substantial departure from the null distribution, indicating that the ETWAS captured a substantial proportion of the transcriptome variability.

### ETWAS Identifies New BD Associations

#### ETWAS Significant Hits

We applied ETWAS to identify genes associated with BD using summary data comprising 20,352 BD cases and 31,358 controls of European descent. ETWAS identified 34 susceptibility genes associated with BD, comprising 44 total associations after Bonferroni corrections (*P* < 2.55×10^−6^, Figure 3A, Supplementary Table 5). For example, the most significant gene, *NEK4*, associated with BD in three tissues (*P*_*CBG*_ = 1.66×10^−9^, *P*_*ACC*_ = 7.38×10^−9^, *P*_*NAB*_ = 1.88×10^−6^).

**Figure 3.**
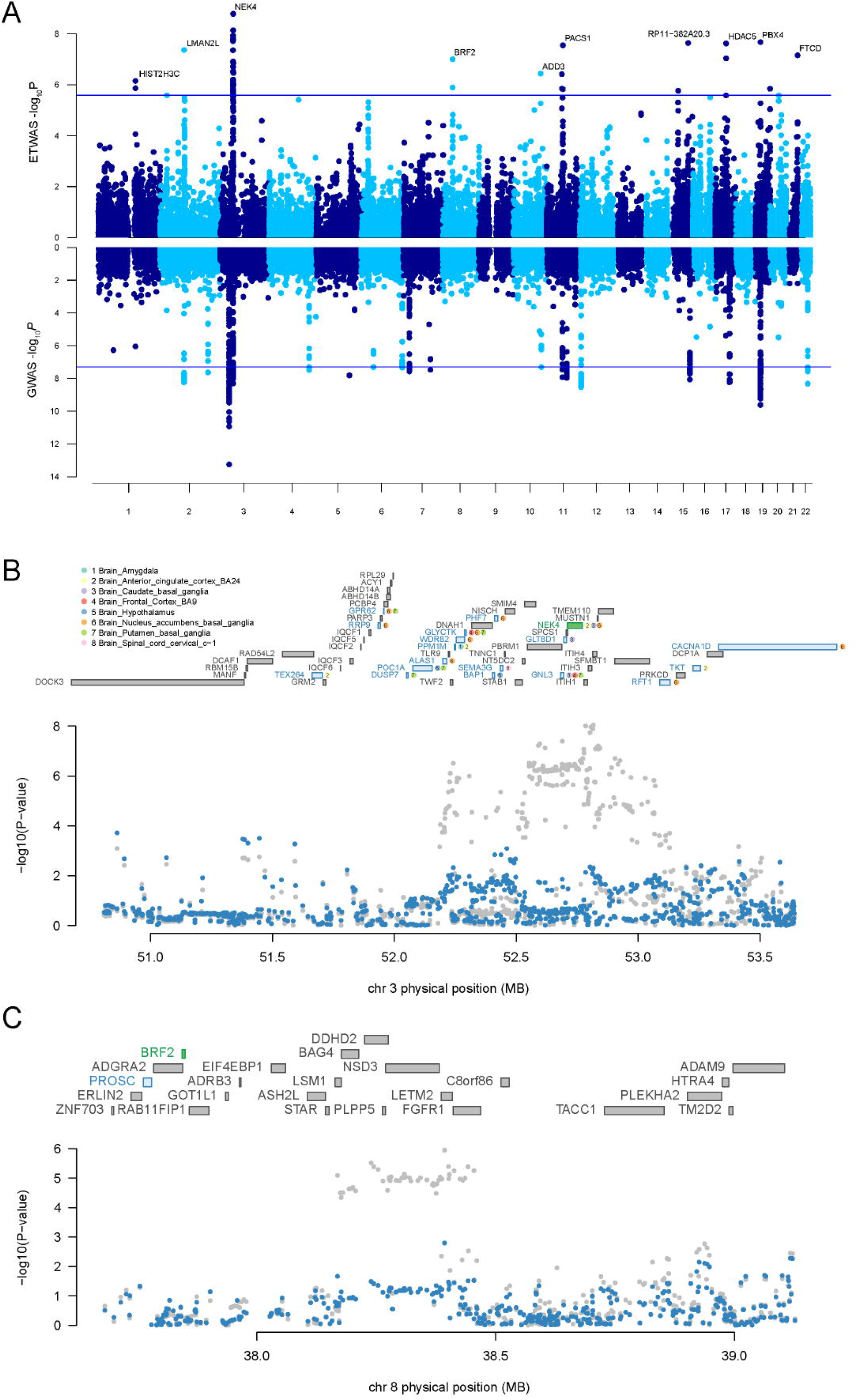
ETWAS identifies new BD associations. (A) Manhattan plot of the ETWAS (upper) and GWAS (lower) results for BD (n = 20,352 cases and n = 31,358 controls). The blue line represents the Bonferroni-corrected significant thresholds, *P* = 2.55× 10^−6^ for ETWAS and *P* = 5×10^−8^ for GWAS. (B-C) Regional association of ETWAS hits. Chromosome 3 (B) and chromosome 8 (C) regional association plot. The marginally associated ETWAS genes are shown in blue and the conditionally significant genes are shown in green. The bottom panel shows a regional Manhattan plot of the GWAS data before (gray) and after (blue) conditioning on the predicted expression of the green genes.

#### Expression signals explain several BD loci

Since several of the ETWAS hits overlapped with significant BD loci, conditional and joint analyses were performed to establish whether these signals were due to multiple-associated features or conditionally independent. We identified 14 conditionally independent genes through conditional analyses. We observed that *NEK4* explains most of the signal at its loci, a region contained 27 significantly ETWAS hits (rs2071044 lead SNP_GWAS_ *P* = 9.10×10^−9^, conditioned on *NEK4* lead SNP_GWAS_ *P* = 0.57, explaining 0.901 of the variances) (Figure 3B). Similarly, conditioning on *PBX4* explains most of the loci’s variance on chromosome 19 (rs1064395 lead SNP_GWAS_ *P* = 9.70×10^−10^, conditioned on *PBX4* lead SNP_GWAS_ *P* = 0.011, explaining 0.583 of the variances) (Supplementary Figure 9A). We also found that *LMAN2L* explains most of the signal at its loci (rs6746896 lead SNP_GWAS_ *P* = 7.00×10^−9^, conditioned on *LMAN2L* lead SNP_GWAS_ *P* = 0.059, explaining 0.674 of the variances) (Supplementary Figure 9B).

#### Expression signals drive BD ETWAS loci

Among the conditionally independent genes, we identified 11 genes that were not implicated in the original BD GWAS, which is regarded as novel candidate genes for BD (Table 1). Conditioning on the expression of *BRF2* explains 0.352 of the variances (rs12677998 lead SNP_GWAS_ *P* = 1.10×10^−6^, conditioned on *BRF2* lead SNP_GWAS_ *P* = 1.60×10^−3^) (Figure 3C). Similarly, conditioning on the expression of *ADD3* in chromosome 10, *FADS1* in chromosome 11, *CDAN1* in chromosome 15, *HDAC5* and *ASB16* in chromosome 17 explains most of the variances of the lead SNP (Supplementary Figure 10).

**Table 1.** Conditionally independent ETWAS genes for BD.

| Gene | Tissue | Best eQTL | <sup>a</sup>Cross-validation R<sup>2</sup> | TWAS Z-score | <sup>b</sup>TWAS P-value | <sup>c</sup>Implicated in 2018 BD GWAS |
| --- | --- | --- | --- | --- | --- | --- |
| NEK4 | CBG | rs2019065 | 0.108 | 6.03 | 1.66×10<sup>-9</sup> | Yes |
| NEK4 | ACC | rs2255107 | 0.073 | 5.78 | 7.38×10<sup>-9</sup> | Yes |
| NEK4 | NAB | rs731831 | 0.131 | 4.77 | 1.88×10<sup>-6</sup> | Yes |
| LMAN2L | SUB | rs11891926 | 0.146 | -5.48 | 4.36×10<sup>-8</sup> | Yes |
| PBX4 | CER | rs2288865 | 0.072 | 5.60 | 2.13×10<sup>-8</sup> | Yes |
| RP11-382A20.3 | CEH | rs8034801 | 0.094 | 5.59 | 2.32×10<sup>-8</sup> | No |
| HDAC5 | CEH | rs7207464 | 0.093 | 5.58 | 2.41×10<sup>-8</sup> | No |
| PACS1 | FRO | rs7114014 | 0.107 | 5.55 | 2.84×10<sup>-8</sup> | No |
| FTCD | NAB | rs2839258 | 0.195 | -5.39 | 7.08×10<sup>-8</sup> | No |
| ASB16 | CEH | rs9910055 | 0.185 | 5.34 | 9.29×10<sup>-8</sup> | No |
| BRF2 | HYP | rs12549353 | 0.141 | 5.33 | 1.01×10<sup>-7</sup> | No |
| ADD3 | CER | rs4918489 | 0.155 | 5.09 | 3.68×10<sup>-7</sup> | No |
| FADS1 | CEH | rs174568 | 0.197 | -5.07 | 3.90×10<sup>-7</sup> | No |
| FADS1 | CER | rs174535 | 0.174 | -4.82 | 1.43×10<sup>-6</sup> | No |
| HIST2H2AA3 | COR | rs2039800 | 0.034 | -4.83 | 1.39×10<sup>-6</sup> | No |
| ZNF584 | SCC | rs1550813 | 0.168 | 4.82 | 1.45×10<sup>-6</sup> | No |
| CDAN1 | CER | rs1359003 | 0.073 | 4.78 | 1.73×10<sup>-6</sup> | No |

<sup>a</sup>The cross-validation R<sup>2</sup> between predicted and observed gene expression is based on tenfold cross-validation within training data. <sup>b</sup>To account for multiple testing, we used a significance threshold of 2.55×10<sup>-6</sup> for all phenotypes. <sup>c</sup>Whether there are any genome-wide significant SNPs within 500 kb away from the gene. Note: CBG, Brain Caudate basal ganglia; ACC, Brain Anterior cingulate cortex BA24; NAB, Brain Nucleus accumbens basal ganglia; CER, Brain Cerebellum; CEH, Brain Cerebellar Hemisphere; FRO, Brain Frontal Cortex BA9; SUB, Brain Substantia nigra; HYP, Brain Hypothalamus; SCC, Brain Spinal cord cervical c-1; COR, Brain Cortex.

### ETWAS increase power to find BD associations

We identified 10 of the 14 conditionally independent genes associated with BD at the gene expression level or the transcript level, including eight novel genes (Supplementary Table 6). Five of the novel genes can be found annotated phenotypes in the knock out mice model. Since the phenotypes MGI arranged did not include BD, we listed all the phenotypes of conditionally independent novel genes reported in MGI (Supplementary Table 7). We found three genes (*HDAC5, ASB16*, and *CDAN1*) associated with cardiovascular disease (CVD) relevant phenotypes, such as cardiac hypertrophy, abnormal heart morphology, abnormal heart shape, and an enlarged heart. There is an integration of the various factors that putatively underlie the association of BD with CVD ^36^, which indirectly suggested that the genes we found may be related to BD.

#### Partitioned Heritability of ETWAS identified BD genes

We partitioned the heritability explained by SNPs around conditionally independent genes and found that conditionally independent genes explained 3.00% (se = 0.46%) of the estimated heritability, an 8.5× enrichment (*P* = 1.31×10^−8^) compared to the percentage of SNPs. The GWAS loci explained 4.58% (se = 0.45%) of the estimated heritability, a 7.9× enrichment (*P* = 5.46×10^−17^) compared to the percentage of SNPs. Combining the SNPs from ETWAS and GWAS explained a much larger percentage of heritability (6.46%, se = 0.65%), a 7.8× enrichment (*P* = 9.29×10^−17^) compared to the percentage of SNPs in either category. We performed t-tests to compare the partitioned heritability from GWAS loci with the partitioned heritability from GWAS loci and the ETWAS identified conditionally independent genes. As shown in Figure 4A, ETWAS identified genes significantly increased the proportion of explained heritability (*P* = 0.019).

**Figure 4.**
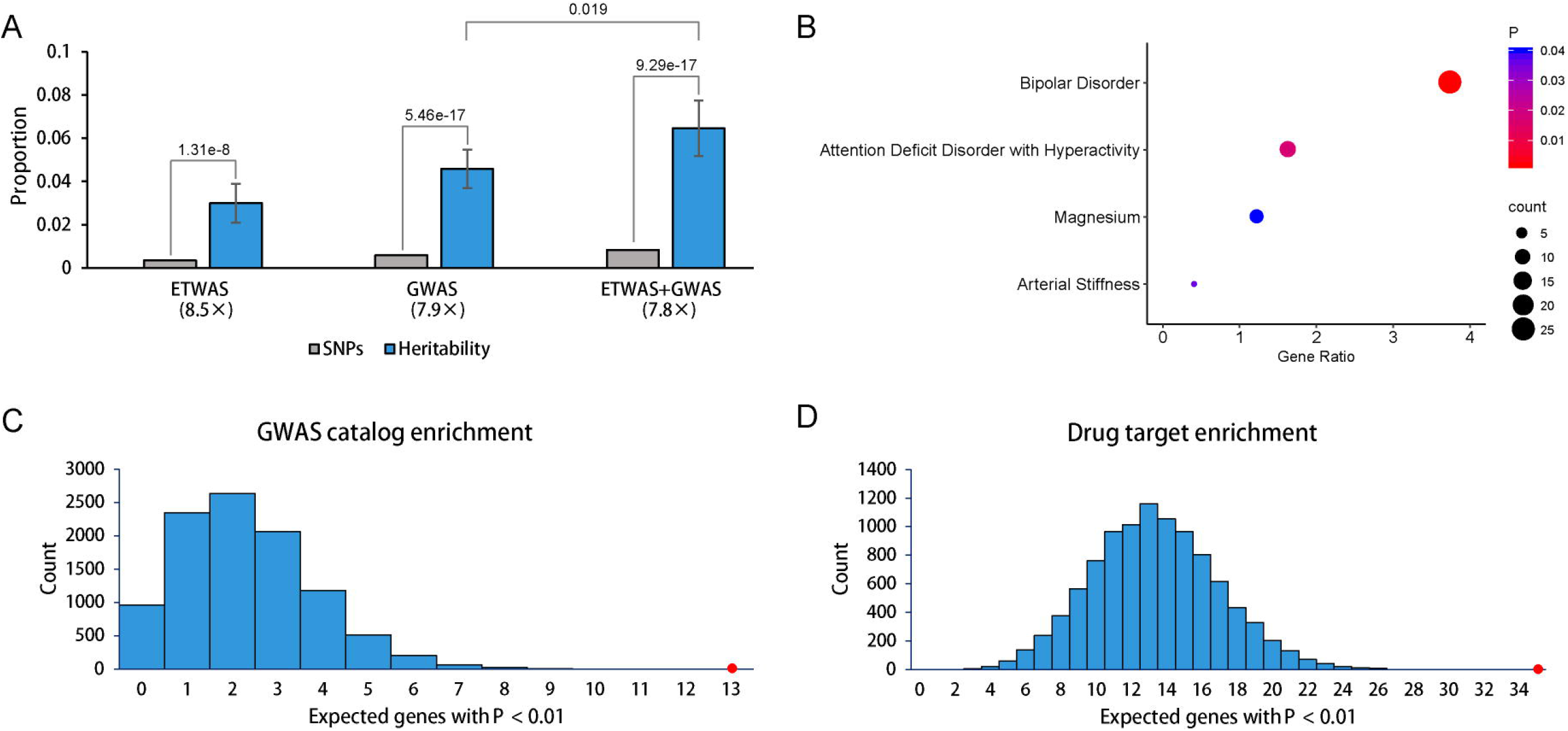
(A) Partitioned heritability of conditionally independent genes and GWAS loci. The null expectation, equal to the percentage of SNPs in each category (gray), and *P* values report the difference from this expectation. Fold enrichment relative to the null expectation is shown in parentheses below each category. Error bars show 1.96× SE. (B) GAD disease enrichment analyses of ETWAS genes. (C) GWAS catalog enrichment analyses of ETWAS genes. The two histograms show the expected number of genes with *P* < 0.01 based on 10,000 random permutations. The large red points show the observed number of previously known BD genes/targets that fall below this threshold. (D) Drug targets enrichment analyses of ETWAS genes.

#### GAD, GWAS Catalog and Drug Target Enrichment Analyses

The GAD disease enrichment analyses detected four significantly enriched diseases (Figure 4B), including bipolar disorder (*P* = 1.7×10^−3^), and attention deficit disorder with hyperactivity (*P* = 0.017). We found that BD had a significant enrichment (*P* < 10^−4^) of GWAS catalog reported genes (Figure 4C) and Open Targets Platform reported drug targets (Figure 4D) in the ETWAS results, which suggested that there are likely to be true disease associations among the genes that fail to meet strict genome-wide significance.

#### Validation of novel loci in subsequent BD study

We further employed ETWAS to identify new expression-trait associations using an early released summary association data for BD in 2012, comprising 7,481 BD cases and 9,250 controls of European descent ^29^. We identified four conditionally independent novel genes using the 2012 BD GWAS summary, that is, associations more than 500 kb away from any genome-wide significant SNPs in that study. We then looked for genome-wide significant SNPs at these loci in the larger 2019 BD GWAS ^4^ (expanded to 20,352 BD cases and 31,358 controls). We identified all the four novel BD-associated genes contained genome-wide significant SNPs in the 2019 GWAS summary data (*P* < 5×10^−8^, Supplementary Table 8). Thus, ETWAS is highly predictive of robust phenotypic associations.

#### Phenome-wide association study

A total of 158 phenotypes, such as cognitive, immunological, metabolic, neurological, psychiatric, were significantly associated with the SNPs in the final model of the BD genes (Supplementary Figure 11A). To determine whether the pheWAS traits were genetically correlated with BD, a genetic correlation was done between BD and the most recent GWAS for each of the phenotypes. We found 38 phenotypes, including nine psychiatric phenotypes, genetically correlated with BD (*P* < 0.05) (Supplementary Figure 11B). After Bonferroni corrections, there were nine phenotypes such as schizophrenia and depression positive correlated with BD. These phenotypes have previously been implicated as risk factors for BD, reaffirming the SNPs’ relevance in the final models.

## Discussion

In this study, we developed a four-step quantitative pipeline named epigenetic element-based transcriptome-wide association studies (ETWAS). We identified 44 genes with genetically-predicted expression levels associated with BD risk. Additionally, through conditional and joint analyses, we identified 14 conditionally independent genes associated with BD risk in 13 brain tissues. Eleven of these genes that not previously implicated with BD, suggesting they are potential novel candidate genes.

We applied conditional and joint association methods to identify genes with significant ETWAS associations when analyzed jointly to account for multiple associations in the same locus. Importantly, the ETWAS expression signals were driving the significance for several previously implicated BD loci when conditioned on the ETWAS genes. For example, the conditioning of *NEK4* led to explain 90.1% of the GWAS signal, which suggests that after considering the predicted expression signal of *NEK4*, there is little residual association signal from the genetic variant in the GWAS locus. We identified 14 conditionally independent genes through conditional analyses, three (*NEK4, LMAN2L*, and *PBX4*) implicated in the original BD GWAS, and the rest eleven genes is regarded as novel candidate genes for BD. Zhihui et al. ^37^ have reported the association between psychiatric risk alleles and mRNA expression of *NEK4*, and the overexpression of *NEK4* could reduce mushroom density spines in rat primary cortical neurons, the most mature form of all spines that responsible for long-term memory. Future studies could interrogate whether expression differences of other candidate genes are consistent with our findings.

Since gene’s heritability providing an upper bound of the predictive accuracy, expression of genes that are not significantly heritable at current sample sizes are not included in this project. Some of the strongly implicated genes in BD risk were not assayed, such as *ITIH1, FADS2*, and *NCAN* ^4^, due to non-significant heritability estimates in any of the brain tissue. We detected the overlap between the conditionally independent genes identified via ETWAS and GWAS reported genes with significantly heritable in at least one brain tissue. We identified only one gene (*LMAN2L*) reported in the GWAS discovery dataset (20,352 cases and 31,358 controls). However, another three genes (*ADD3, HDAC5*, and *PACS1*) were implicated in a more massive combined data with 29,764 BD cases and 169,118 controls (Supplementary Figure 12). Furthermore, two genes (*NEK4* and *FADS1*) were Bonferroni-corrected significant in several brain tissues, while others only significant in specific tissues. Since expression regulation may be common across tissue types, it was refreshing not to see consistency across panels. For instance, *PBX4* had a *P*-value of 2.13×10^−8^ in the cerebellum but a *P*-value of 0.069 in the cortex. Similarly, *HDAC5* had a *P*-value of 2.41×10^−8^ in the cerebellar hemisphere but a *P*-value of 4.90×10^−3^ in the substantia nigra. Although it may be due to tissue-specificity, it is essential to note that it may also be due to specific effects and the quality of the RNA data and panel size of different tissue types from GTEx.

The GAD disease enrichment analyses detected four GAD disease terms, including bipolar disorder, attention deficit disorder with hyperactivity (ADHD), magnesium, and arterial stiffness. Sandra et al. ^38^ conducted a nationwide follow-up study with a total of 2,409,236 individuals investigating the relationship between ADHD and anxiety with the onset of BD. They found that prior diagnoses of ADHD and anxiety disorders are associate with an increased risk of bipolar disorder. By using a Mendelian randomization analysis, Wenwen et al. ^39^ found that magnesium supplementation would increase BD risk. Lithium, the most common treatment for bipolar disorder, could inhibit glycogen synthase kinase-3 by competition for magnesium ^40^. Jess et al. ^41^ performed a cross-sectional metabolic and vascular function evaluation on a sub-sample near completion after a mean follow-up of 27 years and identified chronicity of mood symptoms contribute to vasculopathy in a dose-dependent fashion. GWAS catalog enrichment analyses and drug target enrichment analyses further suggest that ETWAS identified genes are likely to be true disease associations even those fail to meet strict genome-wide significance.

Although we are convinced the method we used to predict gene expression with the combination of genetic variants and regulatory elements has significant potential to delineate further the biological mechanisms for human complex diseases, our current study’s limitations also need to be addressed. First, although gene expression is amenable to genetic prediction with relatively modest sample sizes because of the sparse genetic architecture of gene expression ^35^, recent evidence also suggests that larger expression reference panels will help increase the total number of significant cis-heritable genes available for prediction ^15^. However, due to the limitation of the published transcriptome data sets, the sample size of our reference data was not large enough, as mentioned above. Second, in part due to the historical paucity of eQTL in populations of non-European ancestry, all subjects from the two reference panels were limited to be European ancestry, and the results may not apply to other populations. Since the genetic predictors of gene expression are more accurate in populations of similar ancestry ^42^, further study with a larger sample size of different races with both genotype and gene expression levels is needed. Third, although ultrarare variants have been reported to drive substantial cis heritability of human gene expression ^43^, it is unrealistic to include singletons in expression prediction models at present. Finally, identification of BD-associated genes by ETWAS does not imply causality, and functional studies are needed to determine underlying mechanisms of risk comprehensively. Larger transcriptome and GWAS datasets for BD are likely to improve statistical power for gene identification in the future. Likewise, transcriptome datasets from specific ancestry could also improve future BD ETWAS approaches.

In conclusion, ETWAS is a powerful method that increases statistical power to identify genes associated with BD. We hope ETWAS could provide novel insights into the identification of additional susceptibility genes and further delineate the biological mechanisms for other human complex diseases.

## Data Availability

Raw data available upon request.

