## Supplementary Table 1 for "Epigenetic Element-Based Transcriptome-Wide Association Study Identifies Novel Genes for Bipolar Disorder"

**Supplementary methods**

***Data Resources***

GTEx RNA sequencing data set

In brief, genomic DNA samples obtained from study participants genotyped using Illumina OMNI 2.5M or 5M SNP Array, and RNA samples from 51 tissue sites were sequenced to generate transcriptome profiling data. In this project, we used genotyping and transcriptome data of whole blood and 13 brain tissues to build epigenetic-based gene expression prediction models.

Geuvadis RNA sequencing data set

We obtained freely available RNA-seq data from 358 European lymphoblastoid cell lines produced by the Genetic European Variation in Health and Disease ^1^ (Geuvadis) at <https://www.ebi.ac.uk/>. The genotype data were generated by the 1000 Genomes Project. We used Geuvadis as the validation data set to test the prediction models generated in the GTEx whole blood as the result of cross-study prediction evaluation.

GWAS summary statistics

Cases were required to meet international consensus criteria (DSM-IV or ICD-10) for a lifetime diagnosis of BD established using structured diagnostic instruments.

***Heritability estimation***

We estimated the *cis* heritability (1 Mb window around each gene) for each gene using restricted maximum likelihood analysis, a variance-component model with a genetic relationship matrix (GRM) estimated from genotype data in GCTA software ^2^. A significant nonzero heritability gene defined as genes with 95% confidence not overlapping with zero. Finally, we removed genes in the MHC region and on non-autosomal chromosomes.

***ETWAS Framework***

For each gene, we assessed predictive performance using tenfold cross-validation *R^2^* between predicted and true expression in the initial reference data. We trained and evaluated the models for gene expression prediction in each round of tenfold cross-validation using the following steps. (i) We performed eQTL analyses within 1 Mb of the transcription start/end sites of genes using only the training data to avoid overfitting. (ii) We annotated the eQTL with epigenetic annotations. (iii) We selected multiple SNP sets according to the eQTL *P*-value threshold, chromatin state, TFBS annotation, and DHS annotation. (iv) For each SNP set, we built an expression prediction model in the training dataset by using the lasso and the elastic net (α = 0.5) methods as implemented in the *glmnet* R package.

Second, for each model, we evaluated its prediction performance by the coefficient of determination R^2^ between the predicted gene expression and the observed gene expression of the testing set in fold *y* and averaged all the cross-validation data. For each gene *x*, the model with the highest mean R^2^ was selected as the best model. Based on the parameters of the best model and statistics of eQTL analyses using all samples, we constructed each gene’s final prediction model. Finally, after getting the best model for gene *x*, we could predict expression directly for genotyped samples using the effect sizes from the reference panels and measure the association between predicted expression and a trait. On the other hand, the ImpG-Summary algorithm ^3^ has been used to extend to train on the genetic component of expression based on GWAS summary data ^4^. Thus, we could indirectly estimate the association between predicted expression and a trait as the weighted linear combination of SNP-trait standardized effect sizes while accounting for linkage disequilibrium (LD) among SNPs.

***Partitioned Heritability***

The goal of the partitioned analysis is to quantify the heritability directly explained by SNPs in each functional category using the summary statistics. We partitioned the heritability explained by conditionally independent genes and GWAS loci by SNPs within 500 kb of the genes and GWAS loci. LD score files were made for the two categories separately and simultaneously using the open-source software available at <https://github.com/bulik/ldsc/wiki/Partitioned-Heritability>.

***Enrichment Analyses***

A relaxed threshold of 0.01, rather than Bonferroni-correction, was used for GAD disease enrichment analyses since Bonferroni-correction assumes independence and genes tend to be correlated due to co-expression. More genes will allow for better recapitulation and prioritization of appropriate pathways. GAD disease enrichment analyses were carried out in DAVID (https://david.ncifcrf.gov/home.jsp) using default settings. For the GWAS catalog enrichment analyses, reported BD genes were derived from the comprehensive NHGRI catalog of disease-associated variants identified using GWAS. We excluded studies listed in the GWAS catalog that included the PGC samples to make sure our known gene list was independent of the current analysis. We then counted the number of known disease-associated genes that had an ETWAS P-value below 0.01. We compared this count to the null expectation based on 10,000 randomly drawn gene sets of similar size to the known disease gene set to derive an enrichment P-value. Similarly, we carried out the drug targets enrichment analyses by using the drug targets reported in the Open Targets Platform (https://www.targetvalidation.org/). The Open Targets Platform integrates evidence from genetics, genomics, transcriptomics, drugs, animal models, and scientific literature to score and rank target-disease associations for drug target identification. We consider only targets whose overall association score is higher than 0.1. Similarly, we counted the number of drug targets for BD that had an ETWAS P-value below 0.01 and compared this count to the null expectation based on 10,000 randomly drawn gene sets.

***Differential expression analyses***

The data generated for this analysis represent Freeze 1 and 2 of the PsychENCODE consortium datasets. Details about data generation, RNA-Seq quality control, and normalization were previously reported by Michael et al. ^5^. In brief, several levels of the transcriptomic organization—gene-level, transcript isoform, and local splicing, were generated in brain samples from 1,695 individuals with an autism spectrum disorder, schizophrenia, and bipolar disorder, as well as controls.

***Gene's function analysis based on MGI data***

MGI is the international database resource for the laboratory mouse, providing integrated genetic, genomic, and biological data to assist the study of human health and disease. Knockout mice can be used to identify genes whose deletion results in the disease of mental health.

***Phenome-wide Association Studies***

The top five phenotypes (excluding BD) were reported via public data provided by GWASAtlas (https://atlas.ctglab.nl). To further determine the relationship between BD and the phenotypes identified from pheWAS, genetic correlation of the traits was done for currently available GWAS data via LDSC (https://github.com/bulik/ldsc). The latest GWAS summary statistics were used for the correlation.

**Supplementary Tables**

Supplementary Table 1. Number of samples and significant heritable genes.

| Tissue | | Abbreviation | ^a^Sample Size | ^b^Mean heritability | ^c^Highly heritable genes |
| --- | --- | --- | --- | --- | --- |
| Blood | Whole blood | BLD | 575 | 0.016 | 2,239 |
|  | Lymphoblastoid cell lines | Geuvadis | 358 |  | - |
| Brain | Amygdala | AMY | 120 | 0.043 | 1,192 |
|  | Anterior cingulate cortex BA24 | ACC | 137 | 0.039 | 1,248 |
|  | Caudate basal ganglia | CBG | 174 | 0.035 | 1,573 |
|  | Cerebellar hemisphere | CEH | 159 | 0.046 | 2,010 |
|  | Cerebellum | CER | 190 | 0.046 | 2,606 |
|  | Cortex | COR | 185 | 0.037 | 1,769 |
|  | Frontal cortex BA9 | FRO | 159 | 0.038 | 1,549 |
|  | Hippocampus | HIP | 152 | 0.034 | 1,220 |
|  | Hypothalamus | HYP | 158 | 0.033 | 1,220 |
|  | Nucleus accumbens basal ganglia | NAB | 183 | 0.031 | 1,391 |
|  | Putamen basal ganglia | PBG | 155 | 0.036 | 1,427 |
|  | Spinal cord cervical c-1 | SCC | 116 | 0.047 | 1,335 |
|  | Substantia nigra | SUB | 102 | 0.049 | 1,092 |
|  | Brain Total |  | 1,990 | 0.040 | 19,632 |

^a^Use only European samples; ^b^Average *cis* heritability of all available genes; ^c^The genes located in major histocompatibility complex (MHC) region were removed. The MHC region occurs on chromosome 6, between the flanking genetic markers *MOG* and *COL11A2* (from 6p22.1 to 6p21.3 about 29Mb to 33Mb on the hg38 assembly).

| Type | Source | URLs |
| --- | --- | --- |
| HMM | Roadmap | https://egg2.wustl.edu/roadmap/data/byFileType/chromhmmSegmentations/ChmmModels/coreMarks/jointModel/final/ |
| TFBS | ENCODE | http://hgdownload.cse.ucsc.edu/goldenPath/hg19/encodeDCC/wgEncodeAwgTfbsUniform/ |
| DHS | Roadmap | https://egg2.wustl.edu/roadmap/data/byFileType/peaks/consolidated/narrowPeak/ |

Supplementary Table 2. List of all publicly available epigenomic element files used for training the models.

Supplementary Table 3. Information for the GWASs summary data used in this study.

| Exposure | Case | Control | URL | PMID |
| --- | --- | --- | --- | --- |
| BD_2019 | 20,352 | 31,358 | https://www.med.unc.edu/pgc/download-results/bip/ | 31043756 |
| BD_2012 | 7,481 | 9,250 | https://www.med.unc.edu/pgc/download-results/bip/ | 21926972 |

Supplementary Table 4. The correlation between SNP number in the best model and model performance.

| Tissue | Mean SNP number | Mean R^2^ | r | *P*-value |
| --- | --- | --- | --- | --- |
| Brain Amygdala | 7.70 | 0.13 | -0.10 | 1.48×10^-3^ |
| Brain Anterior cingulate cortex BA24 | 7.82 | 0.13 | -0.08 | 9.73×10^-3^ |
| Brain Caudate basal ganglia | 8.52 | 0.12 | -0.11 | 1.12×10^-5^ |
| Brain Cerebellar Hemisphere | 7.48 | 0.15 | -0.06 | 4.19×10^-3^ |
| Brain Cerebellum | 7.33 | 0.15 | -0.05 | 0.05 |
| Brain Cortex | 7.56 | 0.12 | -0.05 | 0.01 |
| Brain Frontal Cortex BA9 | 8.03 | 0.13 | -0.10 | 2.21×10^-5^ |
| Brain Hippocampus | 8.12 | 0.11 | -0.08 | 1.77×10^-3^ |
| Brain Hypothalamus | 8.14 | 0.11 | -0.09 | 6.88×10^-4^ |
| Brain Nucleus accumbens basal ganglia | 7.84 | 0.11 | -0.07 | 2.91×10^-3^ |
| Brain Putamen basal ganglia | 8.10 | 0.12 | -0.11 | 1.22×10^-8^ |
| Brain Spinal cord cervical c-1 | 8.12 | 0.15 | -0.12 | 9.86×10^-6^ |
| Brain Substantia nigra | 7.84 | 0.16 | -0.13 | 5.93×10^-5^ |

Supplementary Table 5. Significant ETWAS genes for BD.

| Tissue | Gene | Best eQTL | ^a^Cross-validation R^2^ | TWAS Z-score | ^b^TWAS *P*-value |
| --- | --- | --- | --- | --- | --- |
| CBG | NEK4 | rs2019065 | 0.108 | 6.03 | 1.66×10^-9^ |
| ACC | NEK4 | rs2255107 | 0.073 | 5.78 | 7.38×10^-9^ |
| NAB | WDR82 | rs1108842 | 0.098 | 5.70 | 1.20×10^-8^ |
| PBG | POC1A | rs2710323 | 0.064 | 5.70 | 1.22×10^-8^ |
| NAB | PHF7 | rs1108842 | 0.086 | 5.65 | 1.61×10^-8^ |
| NAB | RRP9 | rs13079063 | 0.064 | 5.62 | 1.93×10^-8^ |
| PBG | GPR62 | rs2164884 | 0.097 | 5.62 | 1.94×10^-8^ |
| CER | PBX4 | rs2288865 | 0.072 | 5.60 | 2.13×10^-8^ |
| CEH | RP11-382A20.3 | rs8034801 | 0.094 | 5.59 | 2.32×10^-8^ |
| CEH | HDAC5 | rs7207464 | 0.093 | 5.58 | 2.41×10^-8^ |
| FRO | PACS1 | rs7114014 | 0.107 | 5.55 | 2.84×10^-8^ |
| SUB | LMAN2L | rs11891926 | 0.146 | -5.48 | 4.36×10^-8^ |
| NAB | GPR62 | rs7652667 | 0.064 | 5.47 | 4.46×10^-8^ |
| NAB | FTCD | rs2839258 | 0.195 | -5.39 | 7.08×10^-8^ |
| CEH | ASB16 | rs9910055 | 0.185 | 5.34 | 9.29×10^-8^ |
| HYP | BRF2 | rs12549353 | 0.141 | 5.33 | 1.01×10^-7^ |
| ACC | PPM1M | rs9881468 | 0.104 | 5.29 | 1.25×10^-7^ |
| NAB | RFT1 | rs1004807 | 0.157 | 5.26 | 1.43×10^-7^ |
| HYP | POC1A | rs3755799 | 0.075 | 5.20 | 1.99×10^-7^ |
| FRO | GNL3 | rs7618915 | 0.094 | 5.20 | 2.02×10^-7^ |
| CBG | GLT8D1 | rs8906 | 0.066 | 5.13 | 2.90×10^-7^ |
| PBG | GNL3 | rs3733039 | 0.065 | 5.11 | 3.26×10^-7^ |
| NAB | CACNA1D | rs4282054 | 0.07 | 5.09 | 3.52×10^-7^ |
| CER | ADD3 | rs4918489 | 0.155 | 5.09 | 3.68×10^-7^ |
| CEH | FADS1 | rs174568 | 0.197 | -5.07 | 3.90×10^-7^ |
| NAB | GLYCTK | rs9823697 | 0.101 | 4.98 | 6.25×10^-7^ |
| HIP | HIST2H3C | rs1964542 | 0.067 | -4.96 | 7.12×10^-7^ |
| CBG | GNL3 | rs35526119 | 0.037 | 4.96 | 7.19×10^-7^ |
| SCC | SEMA3G | rs12635140 | 0.041 | 4.93 | 8.40×10^-7^ |
| PBG | DUSP7 | rs10865974 | 0.072 | 4.91 | 9.29×10^-7^ |
| ACC | TKT | rs13071584 | 0.102 | 4.90 | 9.81×10^-7^ |
| ACC | TEX264 | rs12637997 | 0.082 | 4.88 | 1.06×10^-6^ |
| NAB | ALAS1 | rs1080500 | 0.098 | 4.86 | 1.17×10^-6^ |
| HYP | PROSC | rs12335077 | 0.078 | 4.84 | 1.30×10^-6^ |
| COR | HIST2H2AA3 | rs2039800 | 0.034 | -4.83 | 1.39×10^-6^ |
| CER | FADS1 | rs174535 | 0.174 | -4.82 | 1.43×10^-6^ |
| SCC | ZNF584 | rs1550813 | 0.168 | 4.82 | 1.45×10^-6^ |
| AMY | PPM1M | rs11177 | 0.119 | 4.81 | 1.49×10^-6^ |
| FRO | MUS81 | rs1939212 | 0.128 | 4.81 | 1.51×10^-6^ |
| FRO | GLYCTK | rs610060 | 0.1 | 4.79 | 1.65×10^-6^ |
| CER | CDAN1 | rs1359003 | 0.073 | 4.78 | 1.73×10^-6^ |
| NAB | NEK4 | rs731831 | 0.131 | 4.77 | 1.88×10^-6^ |
| HYP | BAP1 | rs2336146 | 0.101 | 4.76 | 1.98×10^-6^ |
| PBG | GLYCTK | rs2564932 | 0.085 | 4.71 | 2.48×10^-6^ |

^a^The cross-validation R^2^ value between predicted and observed gene expression is based on tenfold cross-validation within training data. ^b^To account for multiple testing, we used a significance threshold of 8.41×10^-6^ for all phenotypes. Note: CBG, Brain Caudate basal ganglia; ACC, Brain Anterior cingulate cortex BA24; NAB, Brain Nucleus accumbens basal ganglia; PBG, Brain Putamen basal ganglia; CER, Brain Cerebellum; CEH, Brain Cerebellar Hemisphere; FRO, Brain Frontal Cortex BA9; SUB, Brain Substantia nigra; HYP, Brain Hypothalamus; HIP, Brain Hippocampus; SCC, Brain Spinal cord cervical c-1; COR, Brain Cortex; AMY, Brain Amygdala.

Supplementary Table 6. Differential gene and transcript expression for conditionally independent BD genes in PsychENCODE.

|  | Gene/transcript Name | log2FC/Beta | *P* value |
| --- | --- | --- | --- |
| Differential Gene Expression | BRF2 | 0.054 | 1.50E-03 |
|  | PACS1 | 0.045 | 4.21E-03 |
| Differential Transcript Expression | LMAN2L-008 | 0.422 | 6.81E-05 |
|  | PBX4-006 | 0.247 | 3.33E-03 |
|  | ZNF584-008 | 0.142 | 8.36E-03 |
|  | ADD3-018 | -0.224 | 0.014 |
|  | BRF2-001 | 0.041 | 0.014 |
|  | LMAN2L-002 | -0.197 | 0.018 |
|  | ADD3-021 | 0.183 | 0.023 |
|  | FTCD-006 | 0.132 | 0.027 |
|  | ADD3-003 | 0.048 | 0.035 |
|  | LMAN2L-007 | 0.036 | 0.044 |
|  | PBX4-005 | -0.144 | 0.045 |
| Differential Transcript Usage | FADS1-022 | 0.147 | 2.16E-03 |
|  | PBX4-006 | 2.441 | 6.54E-03 |
|  | PBX4-002 | -1.508 | 0.015 |
|  | HDAC5-007 | -0.325 | 0.017 |
|  | PBX4-003 | -1.186 | 0.040 |
|  | PACS1-001 | 0.872 | 0.043 |
|  | HIST2H2AA3-001 | -1.83E-14 | 0.045 |

Differential Transcript Expression: transcript-level quantifications from RSEM were used as inputs; Differential Transcript Usage: isoform percentage data reported by RSEM was used as inputs.

Supplementary Table 7. Mouse model phenotypes of conditionally independent BD genes.

| Gene | MGI phenotypes |
| --- | --- |
| HDAC5 | cardiac hypertrophy |
| PACS1 | abnormal seminal vesicle morphology |
| ASB16 | abnormal heart morphology; abnormal heart shape; abnormal thymus morphology; decreased circulating alanine transaminase level; decreased circulating alkaline phosphatase level; enlarged thymus; increased caudal vertebrae number |
| FADS1 | abnormal enterocyte proliferation; decreased interferon-gamma secretion; decreased prostaglandin level; decreased tumor necrosis factor secretion; increased prostaglandin level; abnormal circulating lipid level; abnormal lipid level; abnormal mononuclear cell morphology; decreased regulatory T cell number; increased sensitivity to xenobiotic induced morbidity/mortality; premature death; abnormal seminal vesicle morphology; abnormal testis morphology; hypoactivity; improved glucose tolerance; small testis |
| CDAN1 | embryonic lethality between implantation and somite formation, complete penetrance; abnormal embryo size; embryonic growth retardation; abnormal behavior; abnormal heart morphology; abnormal testis morphology; enlarged heart; small testis; embryonic lethality prior to organogenesis; prenatal lethality prior to heart atrial septation; preweaning lethality, incomplete penetrance |

MGI: Mouse Genome Informatics.

Supplementary Table 8. Validation of novel loci in subsequent BD studies.

| Gene | Tissue | ETWAS *P*-value | 2012 GWAS | |  | 2019 GWAS | |
| --- | --- | --- | --- | --- | --- | --- | --- |
|  |  |  | Best SNP | Best SNP *P*-value |  | Best SNP | Best SNP *P*-value |
| TKT | SCC | 4.63×10^-9^ | rs736408 | 2.00×10^-7^ |  | rs2302417 | 4.93×10^-9^ |
| GLT8D1 | NAB | 2.65×10^-8^ | rs736408 | 2.00×10^-7^ |  | rs2302417 | 4.93×10^-9^ |
| GLT8D1 | CBG | 6.96×10^-7^ | rs736408 | 2.00×10^-7^ |  | rs2302417 | 4.93×10^-9^ |
| NEURL3 | SUB | 7.19×10^-7^ | rs6746896 | 4.20×10^-7^ |  | rs57195239 | 5.77×10^-9^ |

Note: Conditionally independent novel genes identified by ETWAS in the 2012 BD GWAS (did not overlap a genome-wide significant SNP in 500 kb), with corresponding best SNP *P*-value in the 2019 study.

**Supplementary figures**

Supplementary Figure 1. The distribution of the number of tissues in which highly heritable genes are located. Among the significantly heritable genes, we found that almost half of them (5,155/9,492) are significant only in one brain tissue.

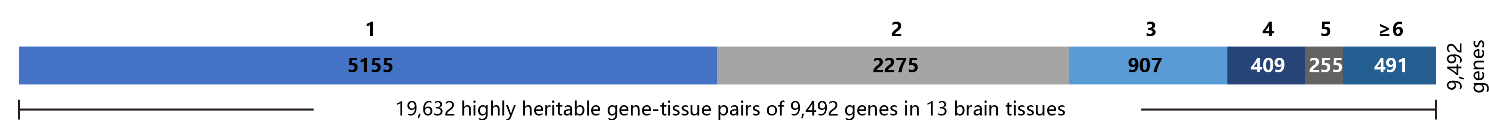

Supplementary Figure 2. Correlation of gene expression heritability in different brain tissues. The correlation is calculated for highly heritable genes with the Pearson method.

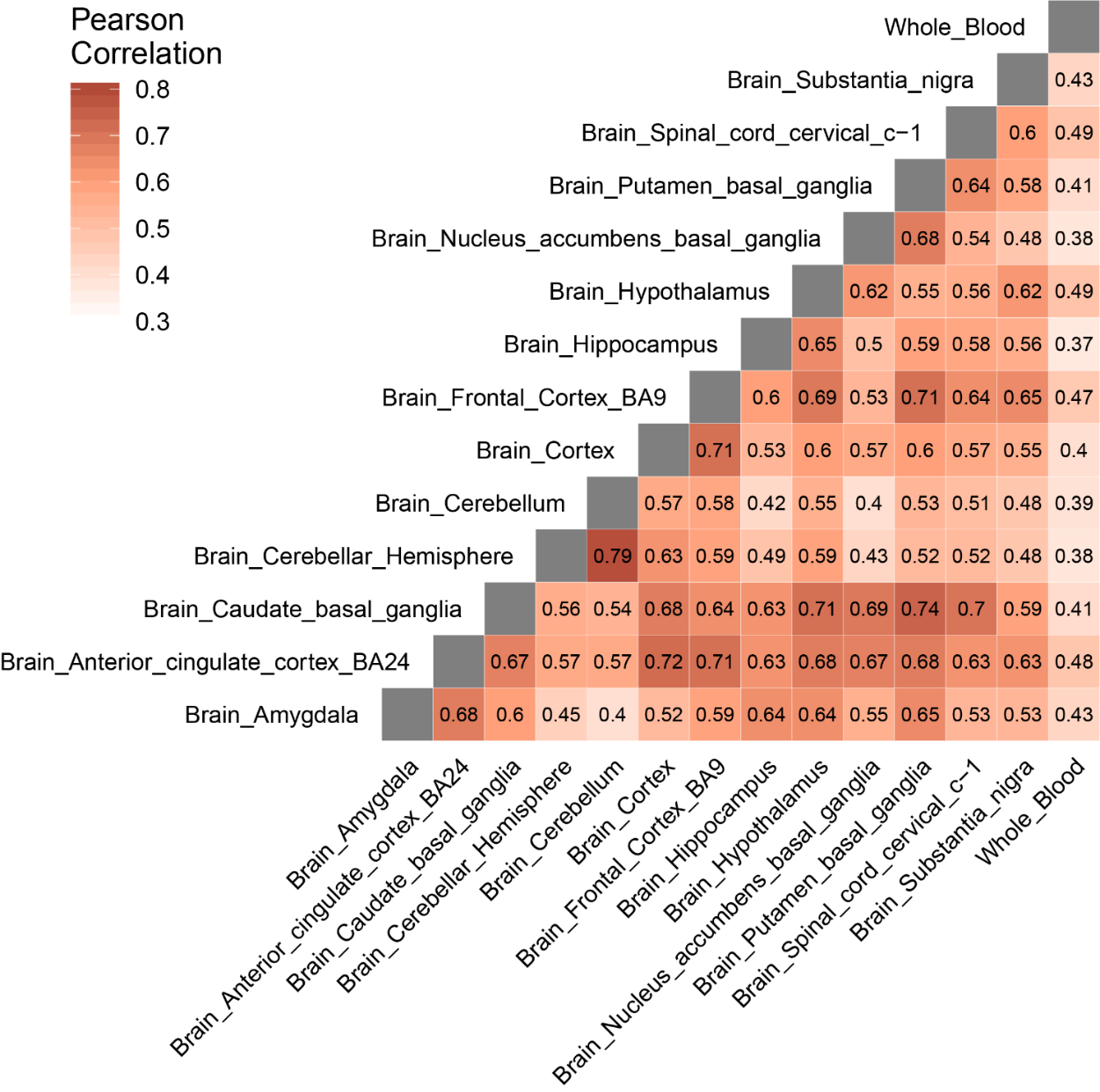

Supplementary Figure 3. Cross-validated prediction performance according to heritability quantiles. The performance of gene expression prediction is better in highly heritable genes.

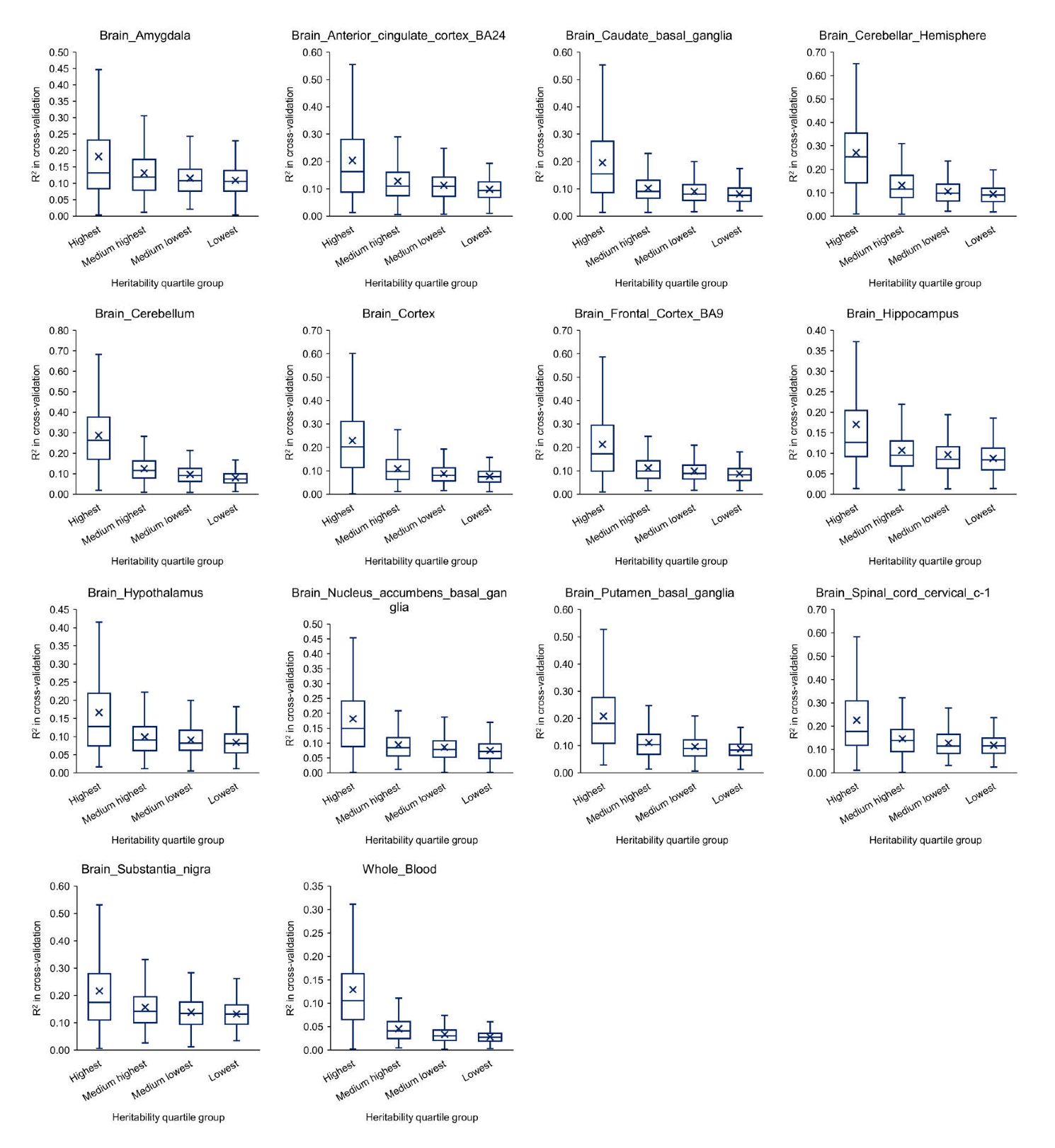

Supplementary Figure 4. The cross-validation R^2^ of the best prediction models in the rest brain tissues were sorted according to the active annotation number and grouped into three categories: 0, 1, ≥2. One asterisk (*) indicates *P*-value smaller than 0.05 (*P* < 0.05), two asterisks (**) indicates *P*-value smaller than 0.01 (*P* < 0.01).

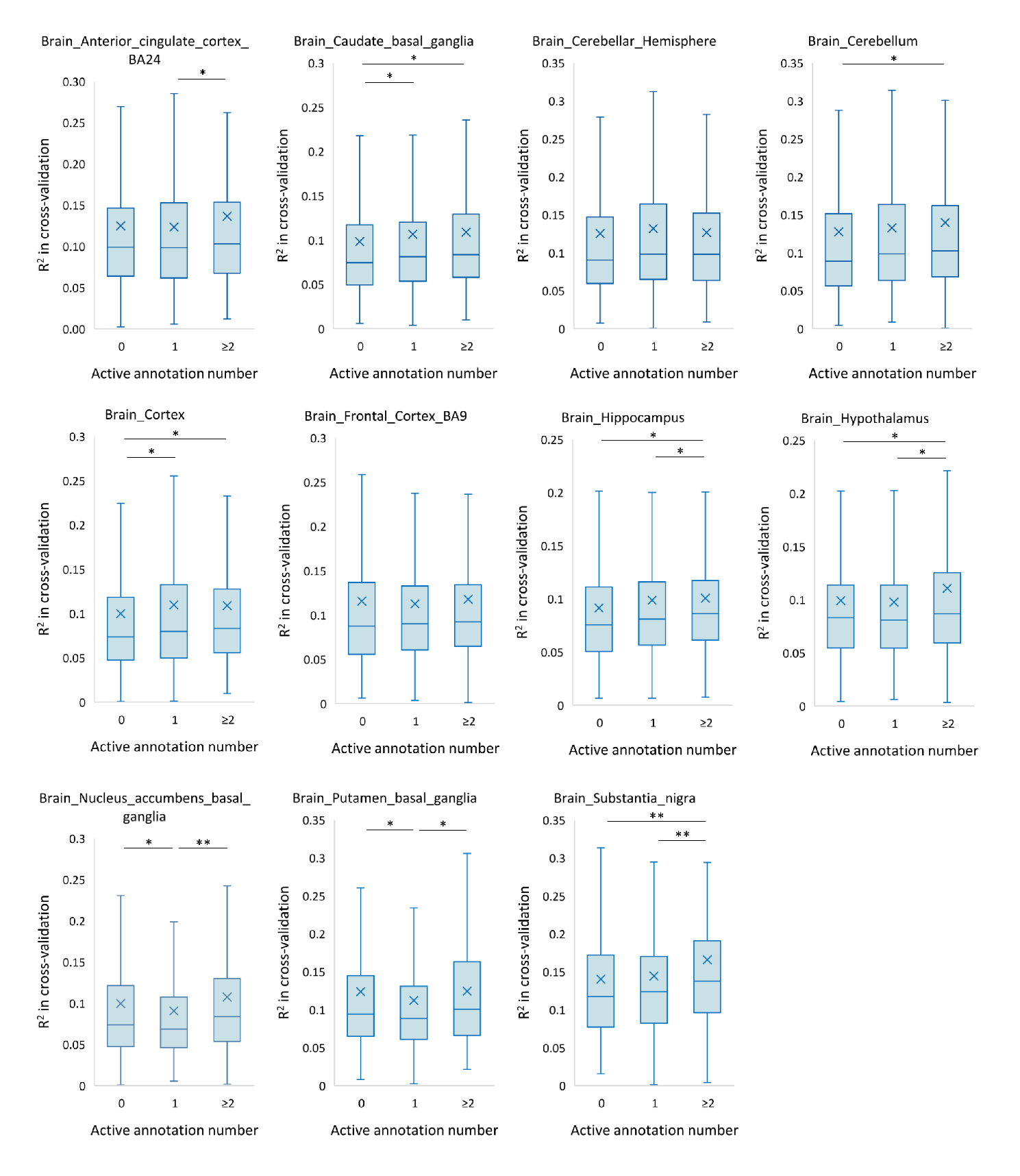

Supplementary Figure 5. The epigenomic annotation distributions of the SNPs used for all genes and the best ETWAS models in brain tissues. (A) The best SNP sets is distributed in the active HMM annotation at a frequency of 1.32-1.45 times that of all variants. (B) The best SNP sets is distributed in the TFBS region at a frequency of 2.12-2.43 times that of all variants. (C) The best SNP sets is distributed in the DHS region at a frequency of 1.70-2.40 times that of all variants.

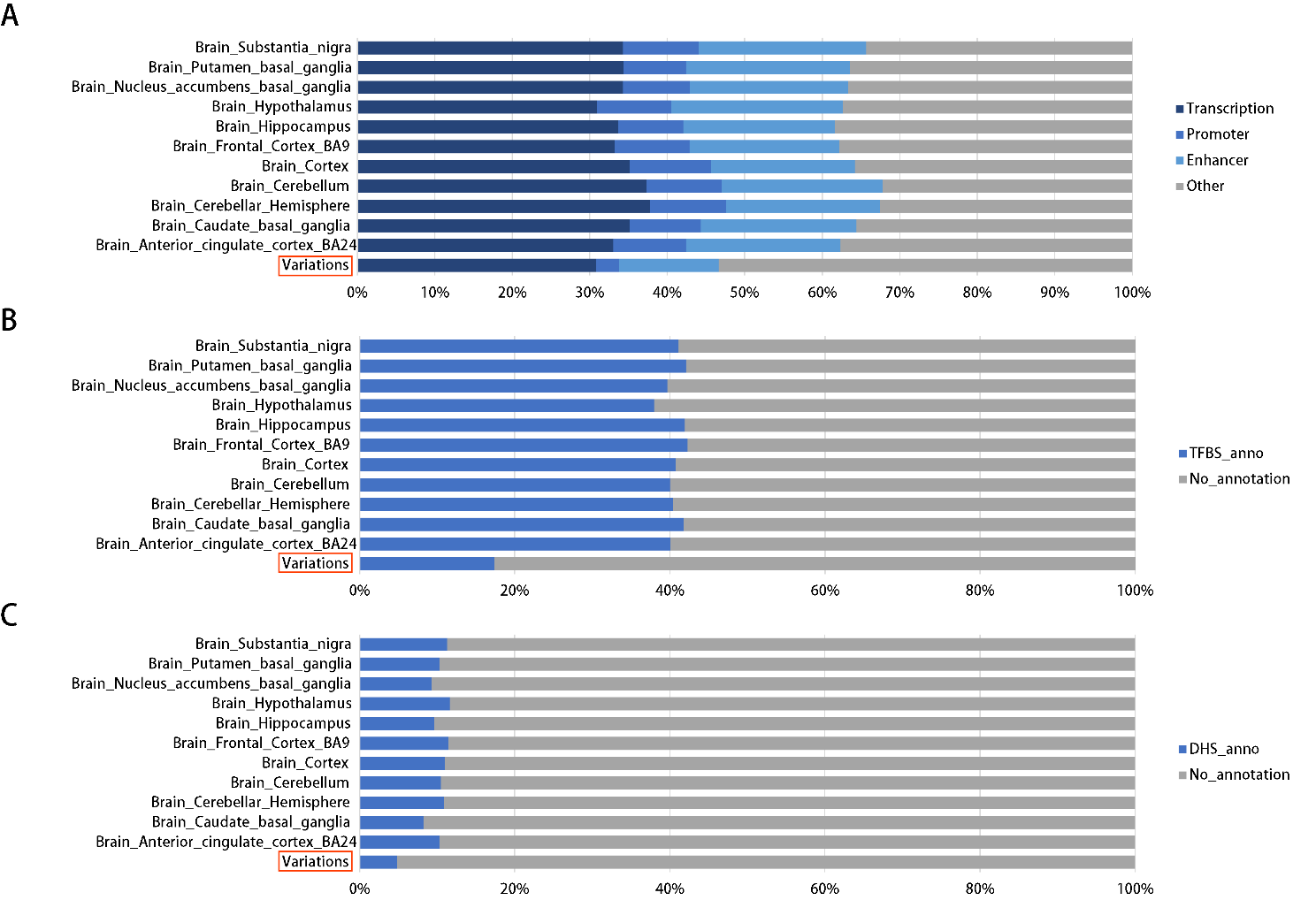

Supplementary Figure 6. Accuracy of individual-level expression imputation models. Accuracy was estimated using cross-validation R^2^ between predicted and true expression. Bars show the mean estimate across 11 cohorts and five methods: Top, single best cis-eQTL in the locus; Lasso, L1 penalty as a variable selection method to select a sparse of predictors; Enet, combines the L1 and L2 penalties of Lasso and ridge regression to perform variable selection; ETWAS.lasso and ETWAS.enet, epigenetic element-based transcriptome-wide association studies (Methods).

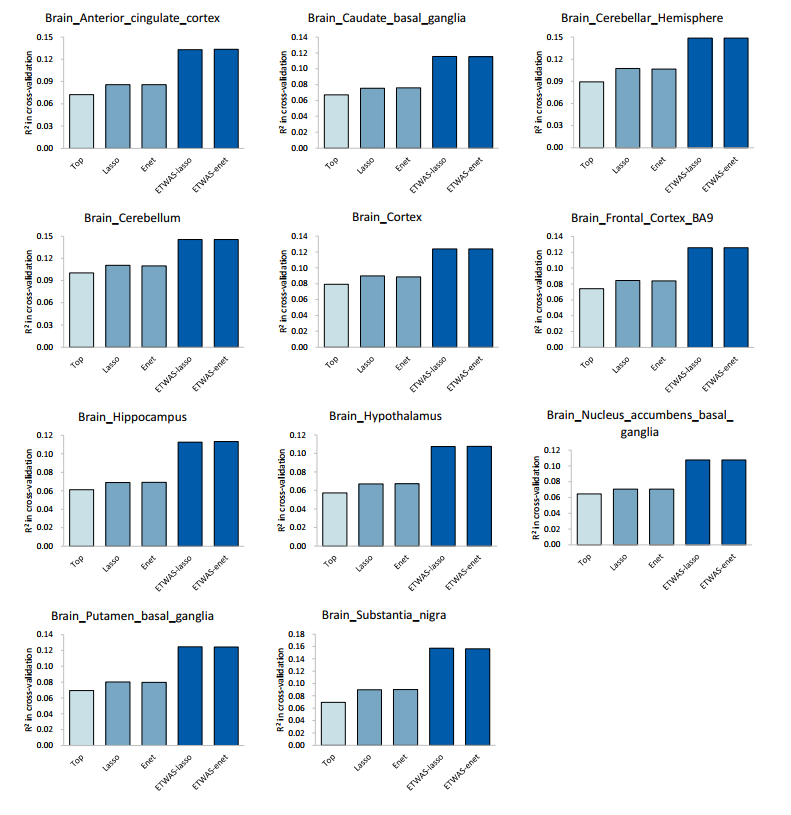

Supplementary Figure 7. Examples of well-predicted genes on a separate cohort. These plots show the observed versus predicted levels of expression for six genes trained in GTEx whole blood. Observed levels were form RNA-seq data in lymphoblastoid cell lines generated by the Geuvadis. Each plot shows the measured expression values (x-axis) versus the expression predictions on the test data (y-axis), where every dot represents one individual in the test data. The liner model (blue line) and 95% confidence interval (gray region) for each plot is also shown along with the gene name, and test R^2^ values.

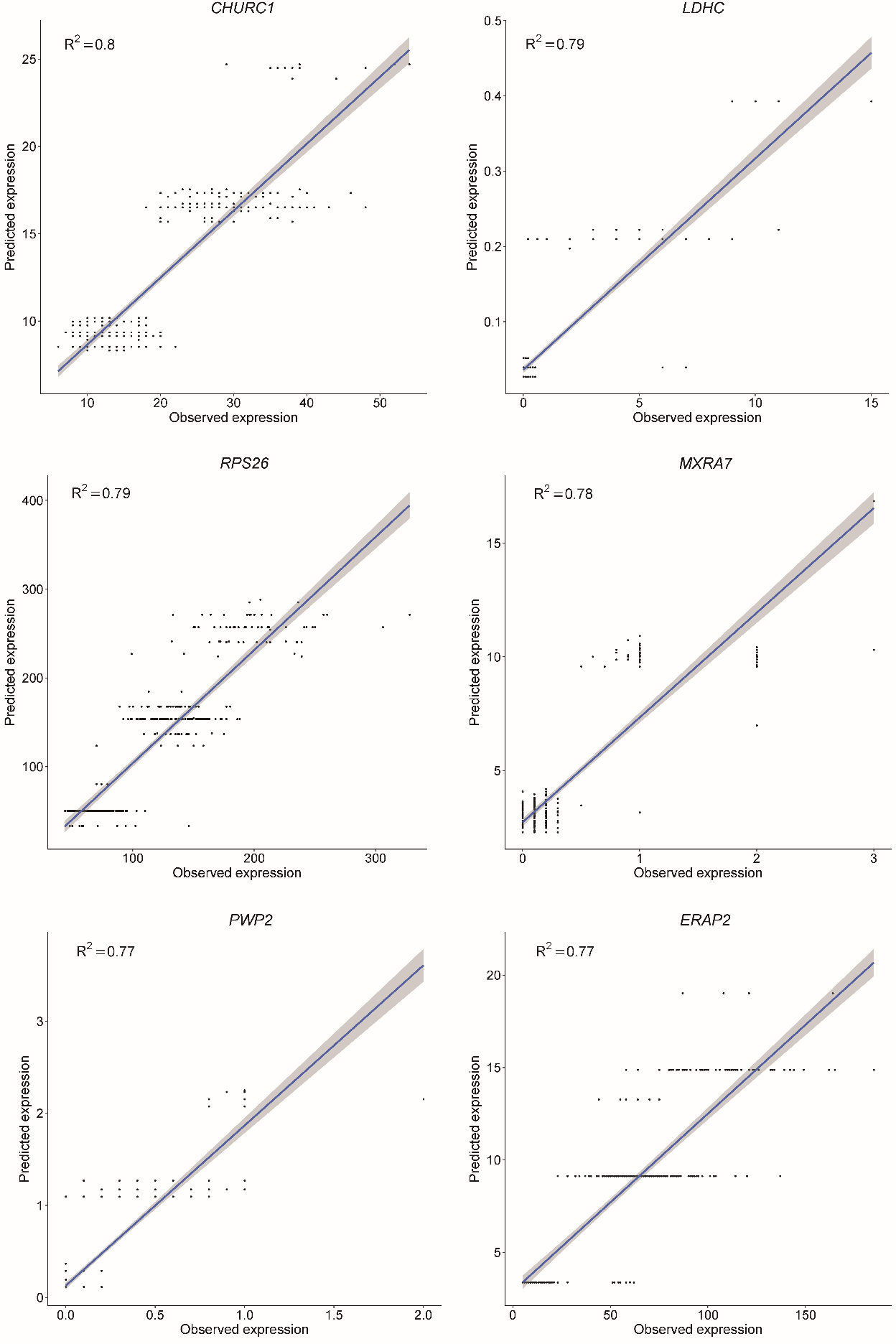

Supplementary Figure 8. Quantile-quantile plot of R^2^ between predicted and observed expression levels in GTEx whole blood.

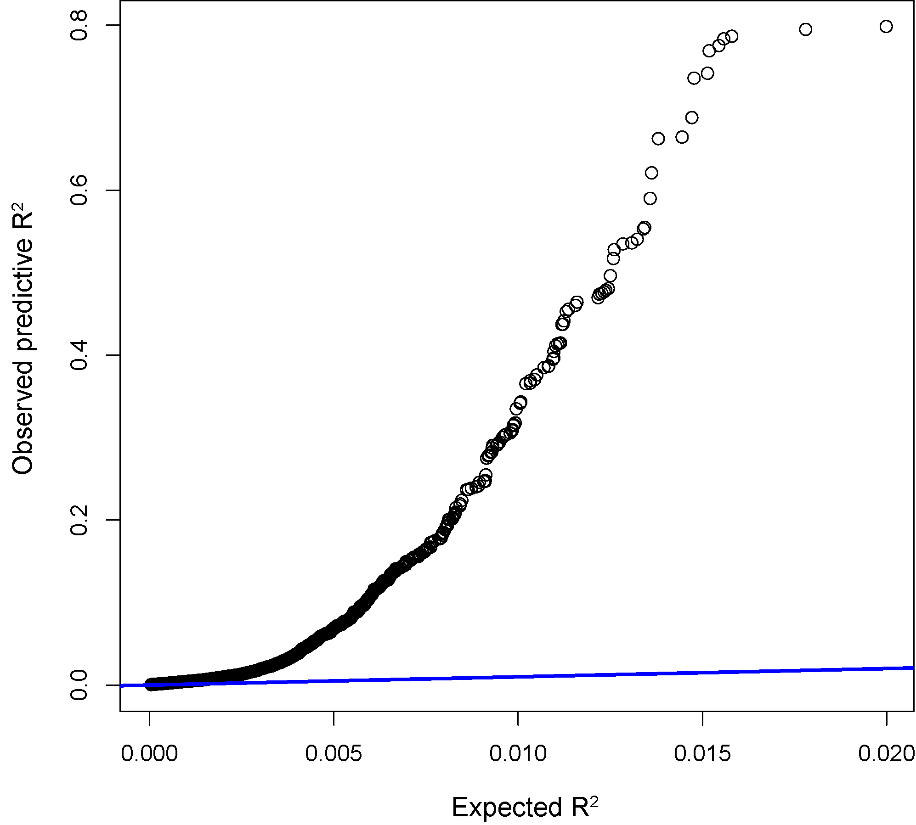

Supplementary Figure 9. Regional association of TWAS hits. The conditionally significant genes are shown in green. The bottom panel shows a regional Manhattan plot of the GWAS data before (gray) and after (blue) conditioning on the predicted expression of the green genes.

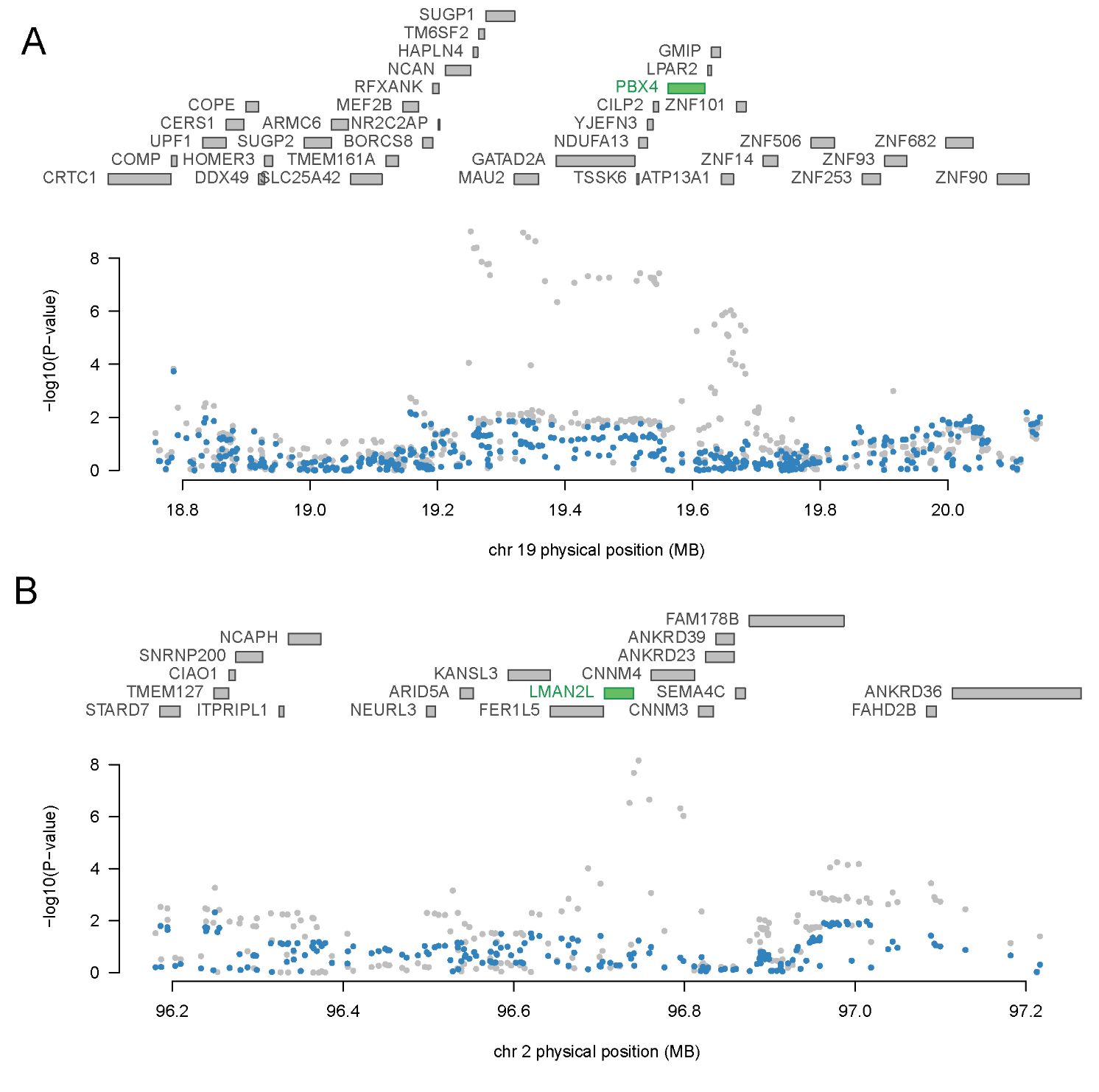

Supplementary Figure 10. Regional association of TWAS hits. The conditionally significant genes are shown in green. The bottom panel shows a regional Manhattan plot of the GWAS data before (gray) and after (blue) conditioning on the predicted expression of the green genes.

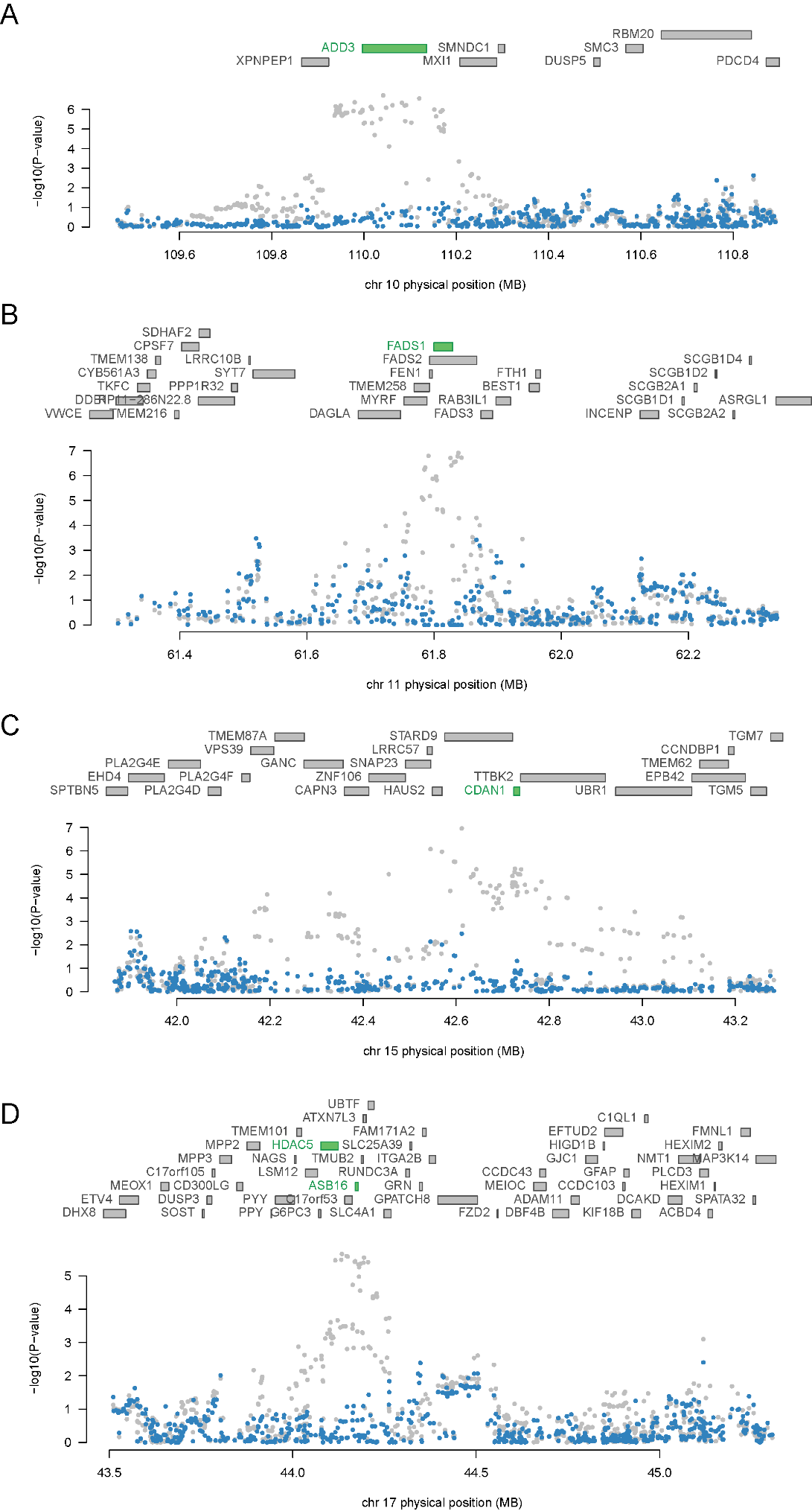

Supplementary Figure 11. (A) Heatmap of the associations between each SNP in the final models of the 14 identified genes and the top five phenotypes (excluding BD) identified via pheWAS. (B) Genetic correlation between BD and phenotypes associated with top BD eQTLs. The asterisk in the box indicates the correlation passes the Bonferroni significance threshold (0.05/158).
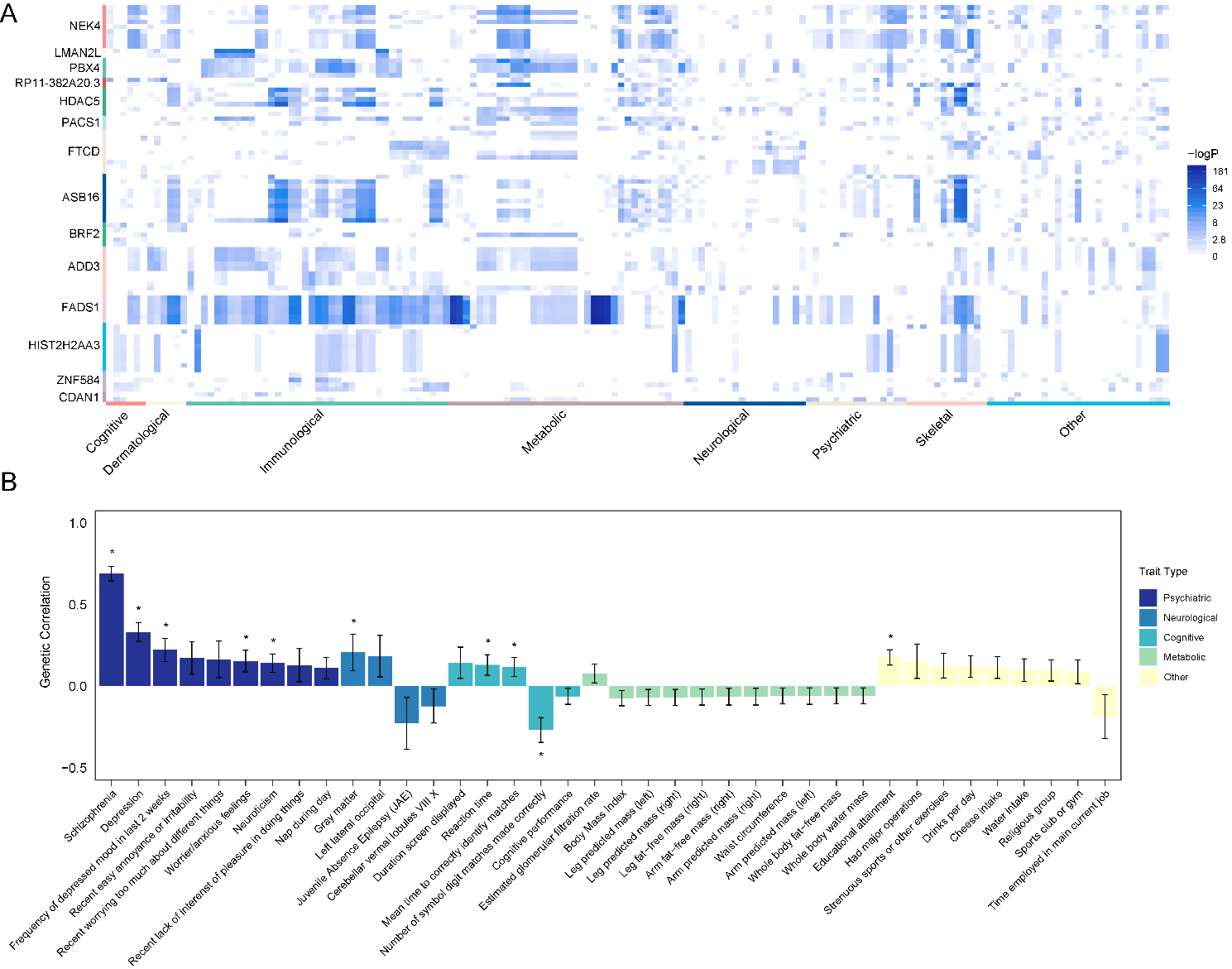

Supplementary Figure 12. The common genes shared by ETWAS and 2019 GWAS data reported genes with highly heritability.

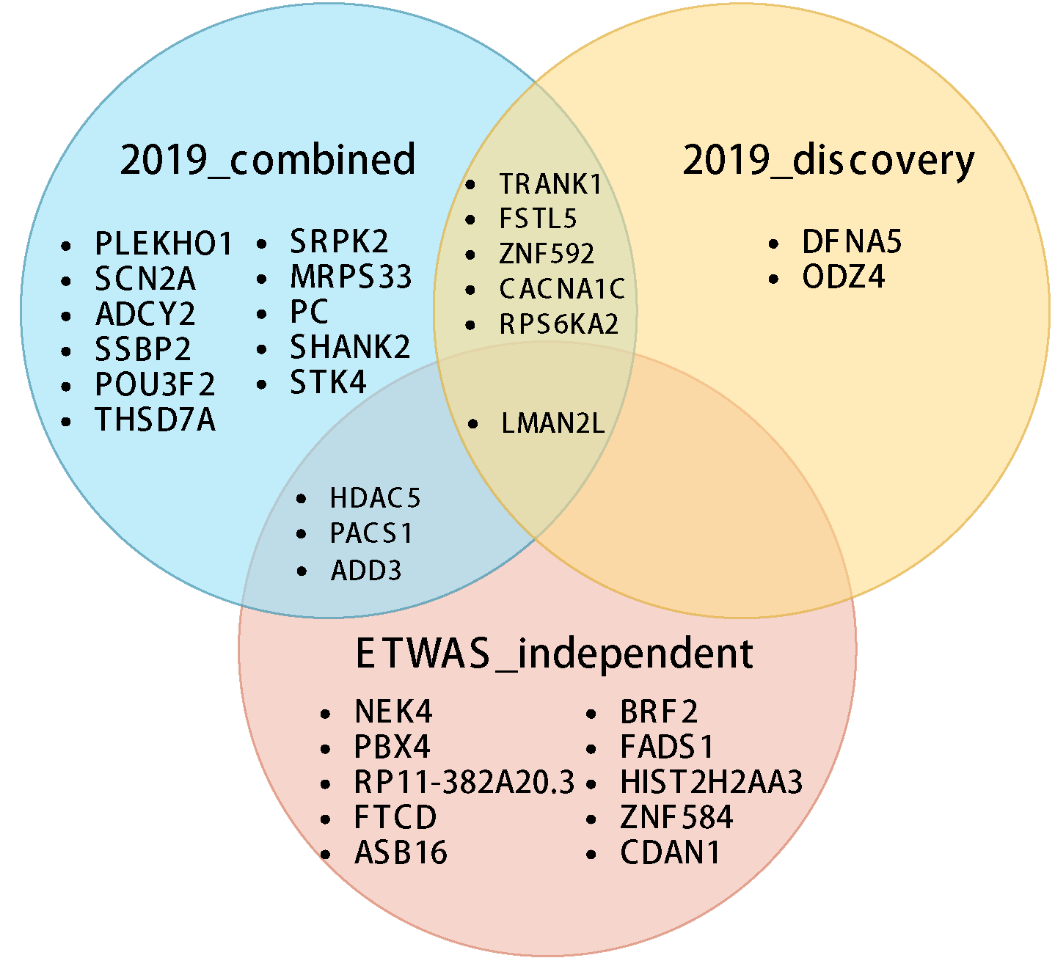
